# Estimating the impact of transfluthrin-treated eave ribbons, Suna traps and their combination on malaria case incidence, based on semi-field and field data

**DOI:** 10.1101/2024.01.29.24301958

**Authors:** Adrian Denz, Margaret M. Njoroge, Mgeni M. Tambwe, Lars Kamber, Aurélien Cavelan, Thomas A. Smith, Joop J.A. van Loon, Alexandra Hiscox, Adam Saddler, Ulrike Fillinger, Sarah J. Moore, Nakul Chitnis

## Abstract

Global malaria incidence has been reduced drastically since the year 2000, primarily due to the widespread use of insecticide-treated nets (ITNs), which kill the bloodfeeding *Anopheles* mosquitoes vectoring the disease in addition to protecting individuals sleeping under them from bites. However, progress has stalled since the mid-2010s and malaria continues to kill more than half a million people globally each year. New, complementary vector control tools are needed to further reduce the residual malaria transmission and face a potential decline in ITN effectiveness due to insecticide resistance. Transfluthrin-treated eave ribbons are a promising spatial repellent to protect people when they are in or around the house but not under an ITN, while the odour-baited Suna trap may present an insecticide-free means of killing mosquitoes. In previous semi-field and field studies, we assessed the effect of the eave ribbon, the Suna trap and the combined push-pull system on mosquito-human contact and mosquito mortality. Here, we combine this evidence and predict the malaria case incidence reduction if these interventions were deployed at a large scale in two East African transmission settings under full uncertainty quantification by use of a stochastic, individual-based simulation platform of malaria epidemiology. Our simulations suggest that the transfluthrin-treated eave ribbon may substantially reduce malaria case incidence in settings with low-transmission or with low ITN use, especially in regions where *Anopheles funestus* dominates among malaria vectors and primarily uses human hosts. However, by diverting mosquitoes from indoor to outdoor host-search, the eave ribbon may reduce the community-protective killing effect of ITNs. In addition, people neither protected by an ITN nor the eave ribbon may experience an increase in malaria incidence at high but imperfect coverage with the eave ribbon. The Suna trap only showed a marginal effect on case incidence and the effect of the combined push-pull system was similar to the effect of the eave ribbon alone. Hence, the eave ribbon appears to be a promising tool in settings difficult to reach with ITNs, such as migrant agricultural workers, but deployment alongside ITNs needs to be planned with care, and ensuring the highest possible use of ITNs remains crucial.

## Introduction

Malaria incidence has been drastically reduced between 2000 and 2015, primarily due to the large-scale implementation of insecticide-treated nets (ITNs)^1^, but progress has stalled since then^2^. Even if coverage, use and formulation of ITNs could be improved further, residual transmission of malaria would still persist, in part because transmission occurs at times and places beyond the reach of ITNs. There is evidence that a substantial part of malaria transmission takes place outdoors and in the evenings; and that the space near houses (peridomestic area), usually within 10 m, where household members spend time before going indoors to sleep, is likely to play an important role in transmission^3–5^. Moreover, two decades of mass ITN use have invoked physiological resistance^6–8^ as well as behavioural adaptations in the mosquito populations^9, 10^ and altered the species composition in many places^11, 12^, rendering *Anopheles funestus* a dominant vector in East Africa^13, 14^. This may further increase residual transmission and jeopardise past achievements, and calls into question a vector control strategy relying mainly on ITNs and indoor residual spraying^15^. Therefore, new tools for mosquito control need to be developed^16–19^.

Spatial repellents are designed to repel mosquitoes from a defined area^20^, but may also prevent host search for an extended time (‘disarming’) or kill mosquitoes depending on the active ingredient and concentrations^21^. Sisal fabrics treated with transfluthrin^22–24^ and fitted to the eave of houses (‘eave ribbon’) showed particular promise to protect against mosquito bites in the peridomestic area^25–27^, especially due to their low cost and minimal need for user compliance. Spatial repellents may, however, push mosquitoes to unprotected humans, increasing their malaria risk^28^. To mitigate this risk, spatial repellents may be combined with odour-baited traps which kill mosquitoes after they are attracted by a synthetic human-odour mimic^29^, similar to ecological push-pull systems in agricultural pest control^30^. Such push-pull systems were shown to reduce house-entry of malaria vectors in the field^31^ and semi-field studies assessed their impact on both indoor biting and outdoor biting in the peridomestic area^32^.

We previously evaluated an eave ribbon treated with 2.5 g emulsified transfluthrin per m^2^ of fabric, the Suna trap^33^ baited with the human odour mimic MB5^34^ as well as carbon dioxide from molasses fermentation^35^ and placed approximately 5 m from the house, and the combined push-pull system in semi-field studies in Mbita, Western Kenya,^27^ and Bagamoyo, coastal Tanzania^36^. Our large semi-field structures of approximately 1000 m^3^ volume allowed for nearly realistic conditions in terms of airflow, temperature and humidity^27^. We showed that both the eave ribbon and the push-pull system strongly reduced biting rates of insectary-reared, plasmodium-free *Anopheles arabiensis* and related these findings to measurements of volatile transfluthrin concentrations in the air^27^, but found no effect of the Suna trap alone in this setting^27^. From these experiments we were able to estimate longer term effects on biting reduction in addition to short-termed repellency^37^, which is important to accurately estimate the impact on malaria transmission. These longer-term effects consist of disarming, defined as preventing host search for one mosquito feeding cycle duration, and killing, but it was not possible to distinguish between the two^37^. In a block-randomised controlled trial over 12 houses and 17 weeks in Ahero, Western Kenya, we tested the same interventions under field conditions^38^, while replacing the carbon dioxide in the trap bait with 2-butanone^39^. In this study, the eave ribbon and the push-pull system strongly reduced indoor biting by *Anopheles arabiensis, Anopheles funestus* and other mosquito species, but had no effect on outdoor biting and even increased outdoor biting by *Anopheles funestus*, presumably due to mosquitoes being diverted from indoor to outdoor biting^38^. The Suna trap again showed no effect on biting rates but caught small numbers of malaria vectors.

Here we combined our field and semi-field findings on the eave ribbon, the Suna trap and the push-pull system by use of a Bayesian evidence synthesis approach, leveraging both the stronger evidence of the field data and the estimates on longer term effects from the semi-field experiments. Based on the obtained estimates of the intervention’s effects on mosquito-human contact and mosquito survival, we then predict their impact on clinical malaria incidence for two baseline settings, we assess deployment strategies with respect to existing vector control interventions, and we present a sensitivity analysis with respect to entomological and epidemiological baseline characteristics. For our predictions we used an open-source, stochastic, individual-based simulation platform of malaria epidemiology^40^ which models the non-linear relationship between malaria transmission and incidence, and allows for detailed calibration to a variety of entomological as well as epidemiological settings. For robustness of our predictions, we propagated the full uncertainty of our entomological effect estimates through the malaria incidence simulations and report Bayesian credible intervals for all outcomes.

## Results

### Differential effects of the eave ribbon, the Suna trap and the push-pull system on malaria transmission

We used a set of five parameters to model the effect of the transfluthrin-treated eave ribbon and the odour-baited Suna trap on mosquito-human contact and mosquito mortality and obtained corresponding Bayesian estimates by combining evidence from our semi-field and field studies (Fig. 1). All interventions showed stronger repellency and biting prevention against *Anopheles funestus* than *Anopheles arabiensis*, due to the latter’s outdoor biting preference and the missing outdoor effect of the eave ribbon^38^. Biting prevention for more than 24 hours (Fig. 1 panel B) concerns the portion of mosquitoes which ceased host-search after exposure to the volatile transfluthrin in the semi-field experiments and thus comprises both disarming and killing. Due to limitations of the semi-field experimental design we could not assess the ratio between disarming and killing, nor the exact duration of the disarming effect.

**Figure 1.**
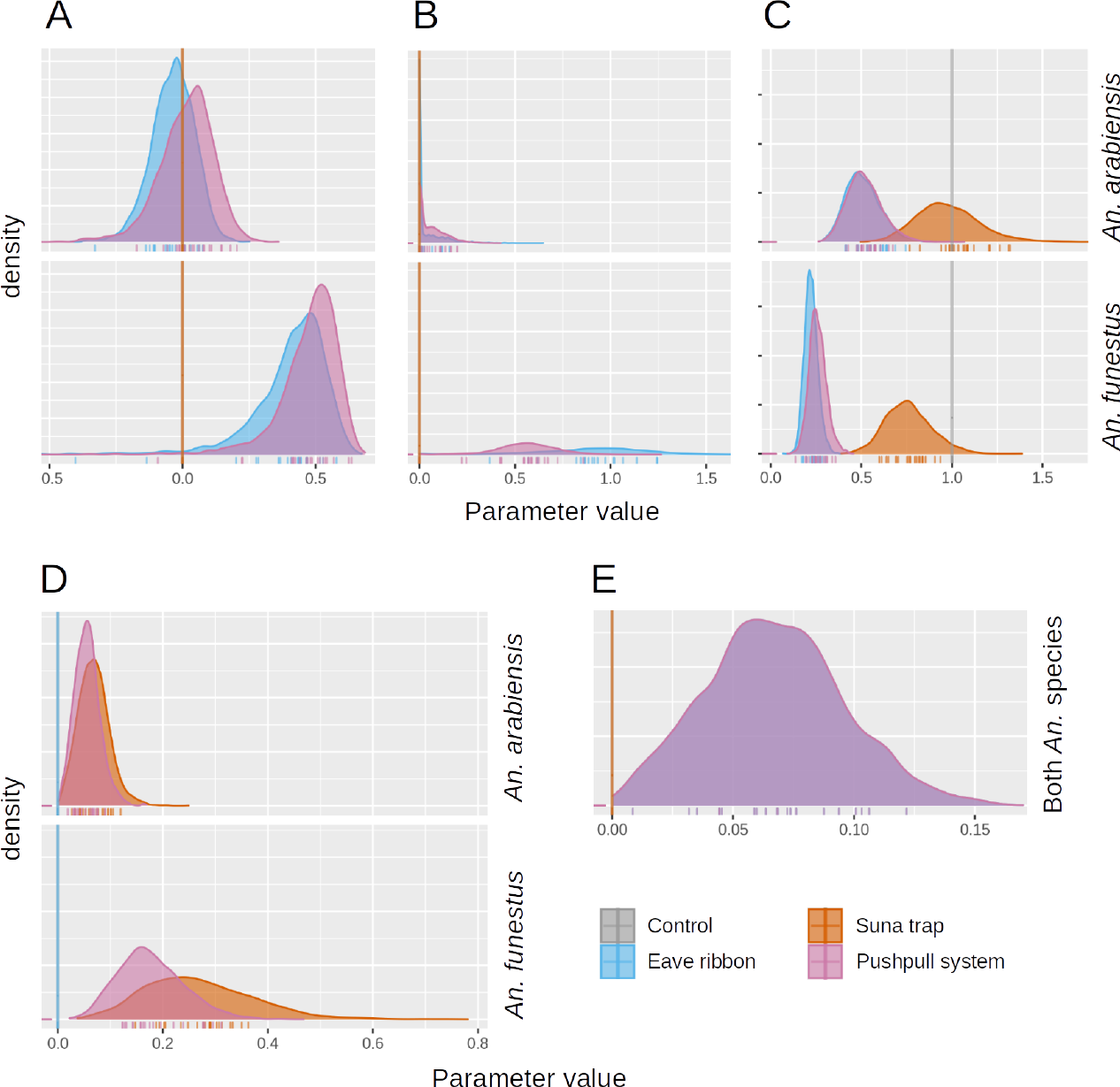
Evidence synthesis of semi-field and field estimated effects of the eave ribbon, the Suna trap and the combined push-pull system on human-mosquito contact and mosquito mortality. All three interventions are modelled by **A** repellency, **B** biting prevention for more than 24 hours, **C** relative change of indoor-outdoor biting ratio, **D** relative attractiveness of the Suna trap compared to an unprotected human outdoors and **E** killing effect after biting, as detailed in 2. For each effect, posterior distributions were computed for the relevant interventions from the semi-field and field data (see Table 1) according to Table 2 by a Bayesian evidence synthesis approach detailed in the methods section. All effects, except killing after biting, are specific for *Anopheles arabiensis* and *Anopheles funestus*. Ticks on the horizontal axes represent the parameter samples used in subsequent simulations of malaria epidemiology.

### Good eave ribbon effectiveness with uncertainty due to assumption on disarming *vs* killing effect

We calibrated an agent-based stochastic simulation platform of malaria epidemiology^40^ according to the epidemiological settings in Ahero, lowland Western Kenya, and the Kilombero valley, South-eastern Tanzania, and simulated the eave ribbon and the Suna trap according to our estimates of the intervention effect parameters while taking into account their full posterior uncertainty (Fig. 1). Our simulations predict that fitting 80% of the houses in a given area with the eave ribbon may reduce malaria incidence by up to 35% if the volatile transfluthrin disarms (prevents biting for 48 hours) and by up to 75% if it kills mosquitoes in addition to repelling them (Fig. 2 panel B). The disarming and killing scenarios concern the portion of mosquitoes which ceased host-search after exposure to the volatile transfluthrin in the semi-field experiments and assume that these either resume host-search after 48 hours or die withing 24 hours, respectively, as their survival couldn’t be assessed in the semi-field experiments^37^. Under the killing assumption, the incremental incidence reduction decreases when increasing coverage of the eave ribbon.

**Table 1.** Data sources for the intervention parameterisation.

| Name | Location and time | Mosquito species | Experimental design | References |
| --- | --- | --- | --- | --- |
| Mbita semi-field data | Mbita, Western Kenya, 2017-2018 | *Anopheles arabiensis* | Semi-field site with parallel control | <sup>27, 37</sup> |
| Bagamoyo semi-field data | Bagamoyo, coastal Tanzania, 2017-2018 | *Anopheles arabiensis* | Semi-field site with parallel control | <sup>36, 37</sup> |
| Field data | Ahero, Western Kenya, 2018-2019 | *Anopheles arabiensis*,<br>*Anopheles funestus* | Latin-square block randomised controlled trial | <sup>38</sup> |

**Table 2.** Intervention parameters for the transfluthrin-treated eave ribbon (R), the Suna trap (T) and the push-pull system (P). If not stated otherwise, parameters are species specific.

| Parameter | Tool | Description | Estimation | Note |
| --- | --- | --- | --- | --- |
| Repellency | R, P | Reduction of availability of protected hosts to mosquitoes, both indoors and outdoors | From field data<sup>38</sup> |  |
| Killing / disarming | R, P | Rate of killing/disarming mosquitoes due to the intervention per protected human | Estimate for *Anopheles arabiensis* from Mbita semi-field data<sup>37</sup> adjusted by ratio of repellency in the field<sup>38</sup> over repellency for *Anopheles arabiensis* in semi-field<sup>37</sup> |  |
| Proportional change of indoor biting | R, T, P | Proportional change of the ratio of indoor biting over outdoor biting due to the intervention | From field data<sup>38</sup> |  |
| Killing after biting | R, P | Relative reduction of the mosquito survival probability after feeding on a protected human | Estimate of R for *Anopheles arabiensis* from Bagamoyo semi-field data<sup>37</sup> (no data available for P) | Not species-specific, uniform for R and P |
| Relative trap availability | T, P | Availability of the Suna trap relative to availability of unprotected human | Estimate of the ratio of Suna trap catch numbers over outdoor biting in the control from the field data, multiplied with proportion of outdoor biting out of total |  |

**Figure 2.**
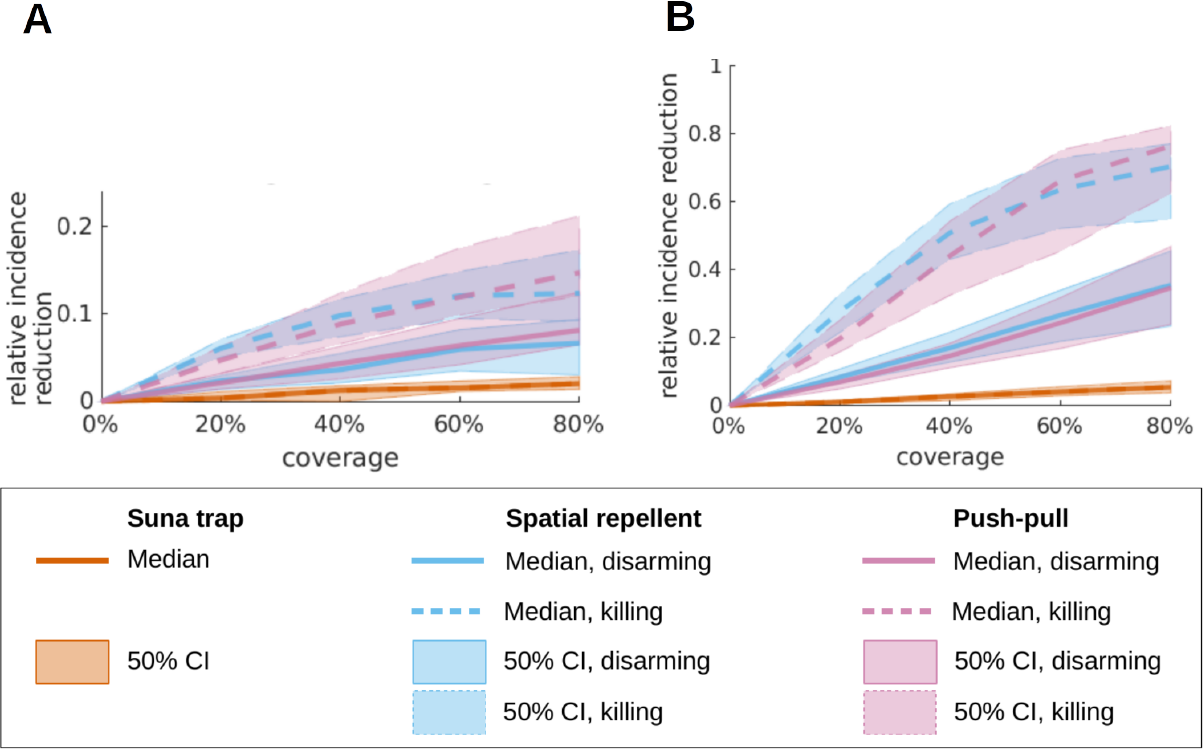
Malaria incidence reduction when increasing coverage with the eave ribbon, the Suna trap or the combined push-pull system. For both settings **A** Ahero and **B** Kilombero, median predictions (lines) and 50% credible intervals (25 to 75% percentile, shaded areas) for relative incidence reduction are reported. The eave ribbon and the push-pull system are simulated under the assumption that the mosquitoes which were prevented from biting for the whole night in the semi-field experiments either rest for 2 days (‘disarming’, solid lines) or die within 24 hours (‘killing’, dashed lines). For each location, a mid-range transmission intensity and ITN coverage of 60% was assumed, corresponding to a baseline entomological inoculation rate of 22 bites in Kilombero and 30 bites in Ahero (Supplementary Fig. S4 and S7). The interventions under consideration are allocated to people regardless of their ITN ownership (random mixing).

### Strong dependence of eave ribbon effectiveness on entomological setting

The effectiveness of the eave ribbon, however, depends strongly on the entomological setting and the transmission intensity as well as to a smaller extent on the baseline ITN coverage level. Baseline incidence was between 1.2 and cases per person and year across all settings considered. In the Kilombero setting, the eave ribbon performs much better compared to the Ahero setting, where the maximal incidence reduction is only about 6% under the disarming assumption and about 12% under the killing assumption (Fig. 2 panel A). In both locations, the dominant malaria vector is *Anopheles funestus*, followed by *Anopheles arabiensis*, while other species have negligible importance^14, 41^. In Kilombero, both *Anopheles funestus* and *Anopheles arabiensis* are more anthropophilic (Human blood index of 100% *vs* 60% for *Anopheles funestus*) and bite more frequently outdoors than in Ahero. This explains the higher effectiveness in Kilombero given the differential effect estimates with respect to the two species (Fig. 1). For increased baseline transmission intensities, relative incidence reduction is lower, about 18% (40% under killing assumption) in Kilombero (Supplementary Fig. S6) and 5% (10% under the killing assumption) in Ahero (Supplementary Fig. S3). Combining the Ahero setting with low baseline transmission intensity, approximating conditions in the Western Kenyan highlands, results in incidence reductions of about 10% and 35% for the disarming and killing assumptions, respectively (Supplementary Fig. S3). When assuming a baseline ITN coverage of 80% as compared to 60%, the eave ribbon is slightly more effective in Kilombero (Supplementary Fig. S6), while there was no difference in Ahero (Supplementary Fig. S3). We modelled testing and treatment of malaria (‘case management’) according to national estimates of effective coverage of the population^42^. Under the assumption of increased case management, both malaria incidence at baseline and the relative effect of the eave ribbon are lower, while sensitivity with respect to the entomological setting as well as ITN coverage is consistent (Supplementary Fig. S5 and S8).

### Marginal effectiveness of the Suna trap and small advantage of the push-pull system over the eave ribbon alone

The Suna trap reduces malaria incidence only marginally in our simulations, with a maximal reduction of 10% if it is deployed at 80% of the houses in Kilombero with a baseline ITN coverage of 80% (Supplementary Fig. S6). Correspondingly, the push-pull system reduces incidence similarly to the eave ribbon alone across all settings under the disarming assumption, while under the killing assumption it is slightly superior at coverage levels exceeding 60% (Supplementary Fig. S3 and S6). Interestingly, the Suna trap performs better in settings with high ITN coverage (Supplementary Fig. S3 and S6).

### Intervention deployment strategy with respect to the baseline ITN intervention

In our simulations, the impact of the eave ribbon and the push-pull system is maximised with a distribution strategy complementing ITN access, meaning that those interventions are first given to people without ITN access (Fig. 3). Once all people in the simulation have access to either ITN or the new intervention, continuing distribution to people with ITN access only has a small effect on averaged incidence across all people in Kilombero (Fig. 3 panel B) and no effect at all in Ahero (Fig. 3 panel A). This antagonism between the eave ribbon and the ITN is due to the eave ribbon pushing mosquitoes away from the house and thus outside of the reach of the killing effect of the ITN. This unfavorable effect is slightly stronger for the eave ribbon alone than for the push-pull system. The small impact of the distribution strategy on trap effectiveness is due to the trap diverting some *Anopheles funestus* mosquitoes from indoor to outdoor biting.

**Figure 3.**
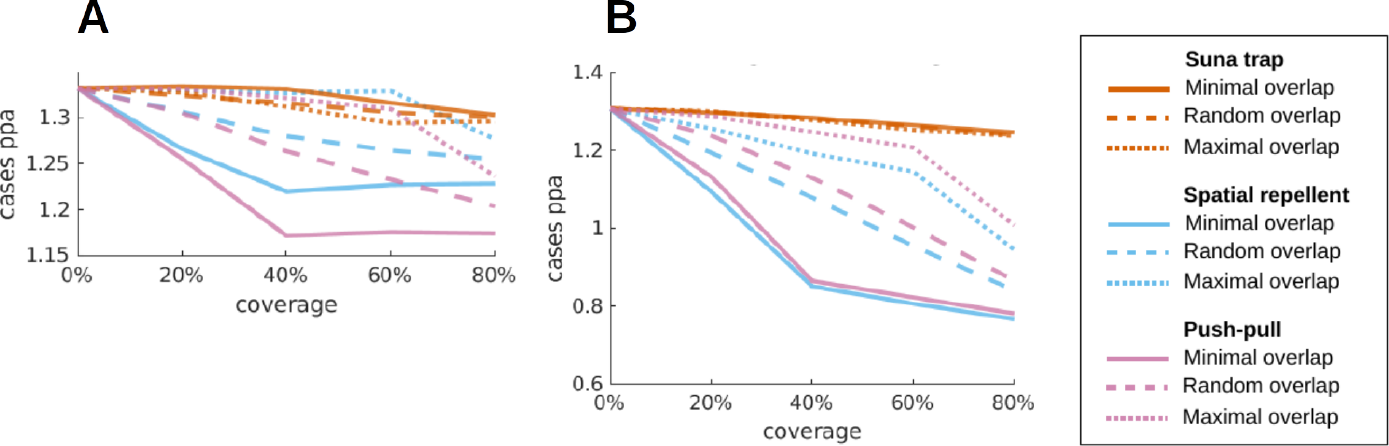
Predicted average malaria incidence for different distribution strategies for the eave ribbon and/or the Suna trap with respect to ITN access. Coverage of simulated people in **A** Ahero and **B** Kilombero with the additional intervention is increased by either serving first people without ITN access (minimal overlap, solid lines), irrespective of ITN access (random overlap, dashed lines) or with ITN access (maximal overlap, dotted lines). All lines denote the median malaria incidence per person and year across all simulations under the disarming assumption. For each location, a mid-range transmission intensity and ITN coverage of 60% was assumed, corresponding to a baseline entomological inoculation rate of 22 bites in Kilombero and 30 bites in Ahero. Similar patterns were obtained under the killing assumption and for different assumptions on the transmission intensity and the ITN coverage.

### Personal *vs* community protection by eave ribbon and push-pull system

We further investigated personal *vs* community protections of the eave ribbon and the push-pull system as well as their interplay with ITNs by tracking incidence per cohort of people with respect to access to ITNs and either of the new interventions. Increasing coverage with the eave ribbon or the push-pull system decreases incidence among people with ITN access in our simulations, irrespective of whether they personally received the new intervention or not (Fig. 4). This community effect is due to the disarming and killing effects, while the diversion from indoor to outdoor biting combined with poor outdoor protection accounts for the missing personal protection. People without ITN access benefit greatly when given the eave ribbon or the push-pull system (Fig. 4). However, for people without ITN access who do not personally receive a new intervention, incidence is unchanged (Ahero, Fig. 4 panel A (c)) or decreases slightly (Kilombero, Fig. 4 panel B (c)) when increasing coverage up to about 40% and rises considerably when increasing coverage further. This unfavorable effect is due to the repelling effect of the eave ribbon which pushes mosquitoes towards unprotected humans in our simulations. This effect aggravates with the number of houses fitted with the ribbon and offsets the community effect by killing or disarming mosquitoes.

**Figure 4.**
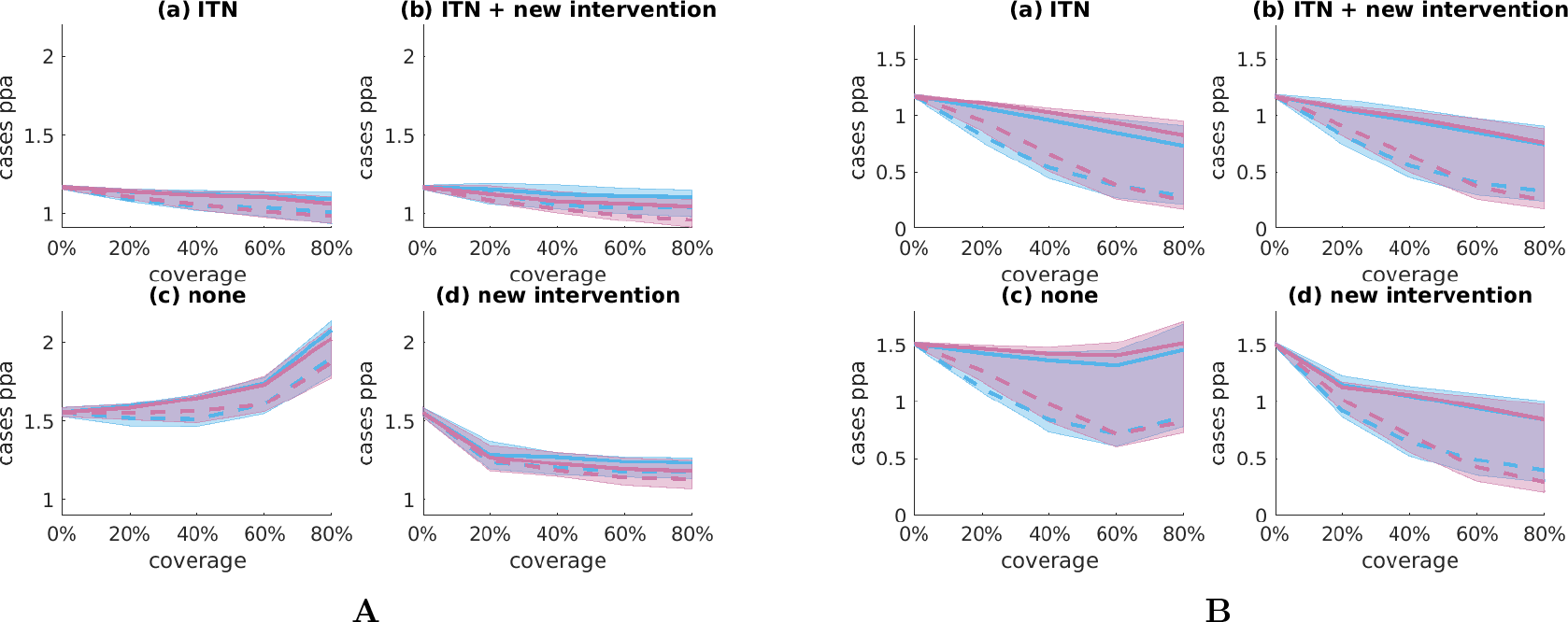
Predicted malaria incidence per cohort with respect to access to ITN and either eave ribbon or push-pull system. For both **A** Ahero and **B** Kilombero, simulated incidence under increasing coverage with the eave ribbon or the push-pull system is reported per group of people with access to either (a) ITN only, (b) ITN and the new intervention, (c) no vector control intervention and (d) the new intervention only. For all simulations, the new interventions are distributed irrespective of ITN access (random overlap). The curves denote the median malaria incidence per person and year across all simulations under either the disarming (solid line) or killing assumption (dashed line). For each location, a mid-range transmission intensity and ITN coverage of 60% was assumed, corresponding to a baseline entomological inoculation rate of 22 bites in Kilombero and 30 bites in Ahero. Similar patterns were obtained under the killing assumption and for different assumptions on the transmission intensity and the ITN coverage. No analysis of personal *vs* community protection was performed for the Suna trap since the trap is not assigned to specific human hosts in the simulation model.

## Discussion

In our model-based predictions, the transfluthrin-treated eave ribbon achieves considerable malaria incidence reduction in transmission settings where *Anopheles funestus* is the dominant disease vector and primarily uses human hosts. Its impact is strongest where transmission intensity and ITN coverage are relatively low. The eave ribbon provides strong personal protection to people without access to ITNs and it provides some indirect community protection due to its disarming and/or killing effect. However, by preventing mosquitoes from entering the house, it diminishes the killing effect of an ITN fitted to the same house, leaving only a small overall community effect. In addition, its repelling effect pushes mosquitoes towards unprotected hosts, which aggravates with increasing coverage and offsets the indirect protection for that group at high coverage levels. Hence, people in our simulations with access to neither ITN nor eave ribbon have an increased risk of getting malaria, which constitutes a violation of the health equity paradigm^43^. Therefore, a targeted intervention strategy for each setting taking into account the local ITN access level and use is required when implementing this spatial repellent. A crossover field study with transfluthrin coils found a strong negative community effect for unprotected neighbours at incomplete coverage (80%) with coils^28^. Similar effects were found for topical repellents^44, 45^.

Contrary to our expectations, the eave ribbon didn’t provide any outdoor protection, potentially due to fast dilution of the transfluthrin by wind^38^. In contrast, previous semi-field^25, 26, 32^ and field studies^25^ with a comparable eave ribbon found a strong outdoor protection effect and even protection of nearby unprotected houses^26^, but in smaller semi-field sites. Basing our study on those data sources would likely have led to quite different results. Interestingly, another modelling study indicated that passive transfluthrin emanators placed further away from the house may push mosquitoes towards indoor host search and thus increase the number of mosquitoes killed by an ITN deployed inside the house^46^. The rationale behind the push-pull system was to mitigate an increase of malaria risk for unprotected neighbours of a house fitted with the eave ribbon. However, the trapping efficiency in both our semi-field and field studies was disappointing, especially in view of previously assumed trapping efficiency^29^, and we see precisely that unfavorable effect in our simulations. Our low trapping efficiency is in contrast to the strong reduction in malaria prevalence during a mass-trapping campaign on Rusinga island, Western Kenya^33^, where the Suna trap was however placed closer to the eave of houses.

Our predictions are based on real data from semi-field and field studies testing the eave ribbon and the Suna trap in Kenya and Tanzania^27, 36, 38^. We accounted for the full uncertainty of the intervention’s incidence impact by sampling all entomological intervention parameters from the corresponding Bayesian posteriors, even when a specific effect was non-significant. This robust approach, together with the uncertainty about the ratio between killing and disarming effects of the eave ribbon^37^, leads to large uncertainty in our predictions. Disarming is a stronger effect than repelling but a weaker effect than killing, especially since it increases the average age and hence the average sporozoite rate of the mosquito population due to the assumed low mortality rate during resting. Hence, the disarming and killing scenarios can be seen as lower and upper bounds of our predictions, respectively.

Our finding of an antagonistic interaction of the eave ribbon with the ITN in terms of community protection is a consequence of diverting mosquitoes from indoor to outdoor biting as estimated from field data^38^. The only relevant modelling assumption here was that diversion happens before a possible interaction with the net, which is needed to deal with the fact that in the field trial there was always an ITN present indoors. In contrast, our findings on increased risk for unprotected people at high coverage levels with the eave ribbon is subject to stronger modelling assumptions. In particular, our transmission model has no notion of space and assumes that repelled mosquitoes can readily encounter any other host. Thus, our predictions of a negative effect for people with access to neither ITN nor eave ribbon may be overly pessimistic.

The predicted malaria incidence reduction potential of the eave ribbon seems especially interesting in view of the expected low unit costs of the product, the minimal technical effort for installation, the low requirements on user compliance and the possibility of local manufacturing^24^. This opens up promising intervention strategies in settings difficult to reach with ITNs, such as migrant agricultural workers^47^. In order to better understand the interaction of the eave ribbon with ITNs as well as indoor residual spraying, experimental huts studies^48^ are needed. Such studies would also allow us to better distinguish between killing and disarming effects. Eventually, larger field trials, preferably cluster-randomised, are needed to assess the impact of transfluthrin-treated eave ribbons on inhabitants of unprotected houses under incomplete coverage. Entomological studies conducted in parallel could clarify a potential effect of the eave ribbon on age structure and sporozoite rate of the mosquito populations. As we believe that climate and weather have a large impact on the effectiveness of the eave ribbon, we suggest to always record weather data. Mosquito control by mass trapping still seems an auspicious strategy due to its insecticide-free mode of action with minimal risk of insensitivity build-up, but the search for the right tool and the right configuration needs to continue. Especially the placement of the trap relative to the house, as indicated by a semi-field study^32^, a practical but sufficient supply of carbon dioxide^49^, and trapping efficiency^50, 51^ may provide room for improvement.

## Conclusion

This modelling study suggests that transfluthrin-treated eave ribbons may substantially decrease clinical malaria in settings with low-transmission or low ITN coverage, especially in regions where *Anopheles funestus* dominates malaria transmission and primarily uses human hosts. People not covered by ITNs benefit from a strong reduction in case incidence when receiving protection by the eave ribbon, opening up promising intervention strategies for settings difficult to reach with ITNs. However, the eave ribbon reduced the killing effectiveness of the ITN and potentially increased the risk for humans still not covered by either the eave ribbon or the ITN. The Suna trap had no significant impact on clinical malaria in the given setup and the push-pull system provided no significant advantage over the eave ribbon. These findings imply that although universal coverage with ITNs needs to remain the major goal of malaria vector control, transfluthrin-treated eave ribbons can play an important role in reducing malaria burden.

## Methods

### Data sources and inference

We used semi-field and field data, as summarised in Table 1, to estimate the parameters used to model the intervention effects in our simulations. For parameter inference, we used Bayesian models which we implemented in the probabilistic programming language Stan^52^ and generated posterior parameter samples with Stan’s default NUTS algorithm^53^ (see Supplementary Methods for details).

### Modelling the transfluthrin-treated eave ribbon

We model three modes of action of the transfluthrin-treated eave ribbon: disarming or killing before encountering a host, repelling, and killing after biting (Table 2). While disarming and killing before host encounter cannot be observed in an open field experiment, we could estimate their combined effect - but not the individual effects – for *Anopheles arabiensis* from the semi-field data^37^. We then adjusted those estimates to the field conditions via the species-specific repelling effects estimated in the field. Disarmed mosquitoes are assumed to rest for the duration of one feeding cycle (2 days) and then restart host-seeking with no further impairments. We model repellency by reducing the availability of protected humans to mosquitoes. Data sources for the effect estimates are given in Table 2; for details on the estimation procedure see the Supplementary Methods.

### Modelling the Suna trap

We modelled the Suna trap as a dummy host which kills all mosquitoes encountering it. We specified the availability of those dummy hosts to mosquitoes, comprising both the attraction and catching efficiency, relative to an unprotected adult’s combined indoor and outdoor availability. Since all humans were protected by a bed net indoors in the field trial, we first estimated the trap’s relative availability with respect to outdoor biting only and then multiplied with the outdoor biting ratio for each species and each setting. For details of the estimation procedure see the Supplementary Methods. It was estimated from the field data^38^ that the Suna trap diverted indoor to outdoor host-search, which we accounted for by altering the indoor biting ratio. The Suna trap is deployed on a household level and we assumed a constant household size of 5 humans for the computation of trap numbers for a given coverage level.

### Modelling the push-pull system

We modelled the push-pull system as an eave ribbon plus a Suna trap, but for each component we estimated its effect by the dedicated push-pull experimental data. In the semi-field experiments, which we used to estimate the disarming or killing effect of the push-pull system, the Suna trap was baited with the human odour mimic MB5^34^ and carbon dioxide produced by molasses fermentation^35^, while in the field experiment, which we used to estimate the other effects of the push-pull system, carbon dioxide was replaced by 2-butanone^39^.

### Modelling the interaction of the transfluthrin-treated eave ribbon with the ITN

ITNs only affect mosquitoes biting indoors and their effectiveness therefore depends on the indoor mosquito density (see the Supplementary Methods for details on the ITN parameterisation). Both the transfluthrin-treated eave ribbon and the Suna trap may prevent house entry of mosquitoes and we therefore modelled their interact with the ITN by modulating the indoor biting ratio according to the proportional change of indoor biting per species which we estimated from the field data (see the for details). For houses where no ITN was present, altering the indoor biting ratio had no effect on transmission in the present model. Furthermore, we assumed that the repellency as well as the killing after biting effects of ITNs and eave ribbons are independent.

### Simulation of clinical outcomes with OpenMalaria

OpenMalaria is an open source simulation platform for malaria epidemiology^54^ which combines a deterministic malaria in mosquito model^55, 56^ with stochastic, individual-based simulations of malaria in humans^57^, including within-host-dynamics, demography and health systems. OpenMalaria models the dynamics of malaria in mosquitoes for multiple species with different characteristics, taking into account seasonality, deployment as well as decay of vector control interventions, and differential infectiousness of humans to mosquitoes. The included mosquito feeding cycle allows for modelling differential intervention effects, but does not distinguish between indoor and outdoor biting explicitly^37^. Hence, we assume that indoor *vs* outdoor biting is a daily behavioural choice of each single mosquito according to a certain species-specific ratio and that infectiousness of mosquitoes is homogeneous within each species. OpenMalaria was calibrated and tested against multiple data sets^58^ and the source code is freely available^40^. All simulations are run with a human population of 10,000 people.

### Simulation setup

We considered 20 baseline settings and 73 intervention scenarios (Fig. 5). For each coupling of a baseline setting with an intervention scenario, we ran 20 OpenMalaria simulations, each with a different random seed to initialise the simulator and a different draw from the posteriors of the intervention parameters estimated from the real data. All simulations start in the year 1916 with a warm-up phase of one maximal human lifetime to ensure the entire human population has a realistic level of natural immunity to malaria (Fig. 6). From 2006 onwards, ITNs are distributed in a three-year cycle through mass campaigns as a baseline for all scenarios, corresponding to the scale up of ITNs in the African region starting approximately at this time. In 2021, the interventions under consideration are deployed and maintained at constant effectiveness until 2024, when the simulations are terminated. Processing the input to OpenMalaria has been performed in R^59^. Postprocessing of the simulated data and visualisation has been performed in MATLAB^60^.

**Figure 5.**
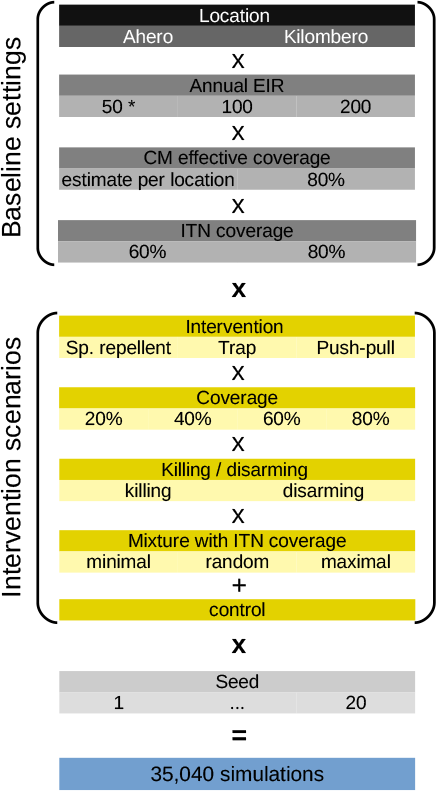
Experimental setup. The seed determines both the seed to initialise the random number generator for the OpenMalaria simulation and the draw from the posterior of the intervention parameterisation. Note that the annual entomological inoculation rate (EIR) specified here determines the transmission intensity before deployment of ITNs. * An annual EIR of 50 is only simulated for Ahero.

**Figure 6.**
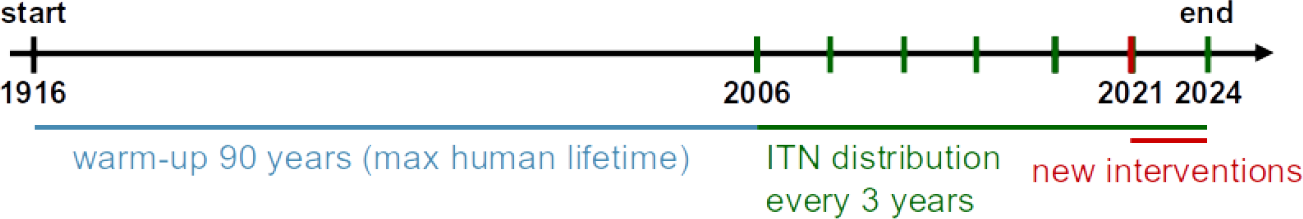
Timeline of OpenMalaria simulations.

### Locations

The two geographic settings we chose, Ahero in Western Kenya and the Kilombero valley in South-eastern Tanzania, differ in their mosquito bionomics, health systems and demographics, as summarised in Table 3. We only modelled *Anopheles arabiensis* and *funestus*, as all other species, including *Anopheles gambiae sensu stricto*, were reported as not abundant or only marginally abundant in the two studied locations^14, 41^. For further assumptions on mosquito bionomics and the seasonality with which the entomological inoculation rate (EIR) varies over the year, see the Supplementary Methods. The age structure of the human population was based on previous parameterisations of Western Kenya^61^ and the Kilombero Valley^62^. The effective coverage (*E*_14_) of case management (CM) with artemisinin-based combination therapy was set to national estimates^42^.

**Table 3.**
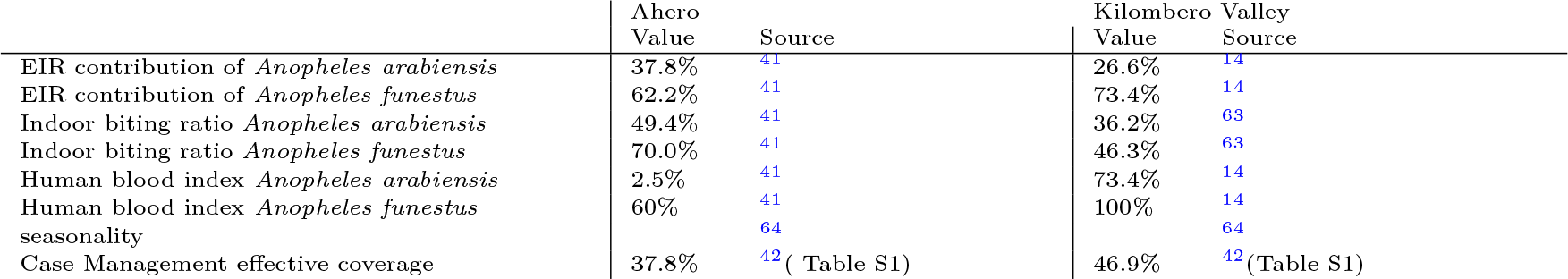
Parameter choices for baseline settings Ahero and Kilombero Valley. EIR refers to the entomological inoculation rate.

|  | Ahero | Source | Kilombero Valley | Source |
| --- | --- | --- | --- | --- |
| EIR contribution of *Anopheles arabiensis* | 37.8% | 41 | 26.6% | 14 |
| EIR contribution of *Anopheles funestus* | 62.2% | 41 | 73.4% | 14 |
| Indoor biting ratio *Anopheles arabiensis* | 49.4% | 41 | 36.2% | 63 |
| Indoor biting ratio *Anopheles funestus* | 70.0% | 41 | 46.3% | 63 |
| Human blood index *Anopheles arabiensis* | 2.5% | 41 | 73.4% | 14 |
| Human blood index *Anopheles funestus* | 60% | 41 | 100% | 14 |
| seasonality |  | 64 |  | 64 |
| Case Management effective coverage | 37.8% | 42 (Table S1) | 46.9% | 42 (Table S1) |

### Sensitivity Analysis per location

#### Transmission intensity

For each location, we simulated mid-range and high transmission intensities, corresponding to EIRs of 100 and 200 as measured before implementation of ITNs but after case management (CM), respectively. For Ahero, we additionally simulated a low transmission setting with an annual EIR of 50 as a proxy of highland settings in Western Kenya. We attributed these EIR levels to the different species according to the species composition per setting (see Table 3).

#### Baseline vector-control interventions

We assume ITNs are distributed by mass campaigns in 3 year cycles from 2006 onwards and we simulated 60% as well as 80% coverage to account for variation in ITN access. ITNs only act on mosquitoes biting indoors and we therefore modulated the ITN effectiveness according to the indoor biting proportion for each species and each location (see Table 3). For details on the ITN parameterisation in OpenMalaria see the Supplementary Methods.

#### Case-Management / health system

In addition to the location specific estimates of the effective CM coverage (see Table 3), we simulated an increase in effective CM coverage to 60% per location. Due to the used EIR definition, two baseline settings with the same transmission intensity and different CM coverage levels must be seen as two distinct entomological settings with the same transmission intensity resulting from different CM coverage levels, and not as a single entomological setting with two CM strategies.

#### Intervention scenarios

The eave ribbons, the Suna trap and the push-pull system are deployed separately with coverage levels of 20, 40, 60 and 80% of the human population. We considered three deployment strategies with respect to ITN access – either serving first people without ITN access (‘minimal overlap’), irrespective of their ITN access (‘random overlap’) or with ITN access (‘maximal overlap’). For the ‘random allocation’ scenario, we reported incidence per cohort receiving none, one or both of the interventions, with at least 400 individuals per cohort. For the eave ribbon and the push-pull system, we considered two distinct scenarios for killing and disarming mosquitoes. We assumed the eave ribbon and Suna trap are replaced as necessary and thus no decay of their effectiveness over time.

## Data Availability

All data produced in the present study are available upon reasonable request to the authors.

## Authors’ contributions

MMN, FO, JvL, AH, AS, UF, SJM and NC designed the experiments. MMN and MMT performed the experiments. AD, NC, TS, LK and AC developed the modelling methodology. AD performed the Bayesian inference. AD ran the simulations and created the figures. AD and NC interpreted the results. AD wrote the initial draft of the manuscript. AD and NC contributed to writing the manuscript.

## Acknowledgements

We thank Clara Champagne for helpful discussions and suggestions, and Naomi Richner for proofreading the manuscript.

## Supplementary information

### Supplementary Methods

#### Estimation of intervention parameters from field and semi-field data

#### Note on differences between field and semi-field data

It is not possible to use the semi-field model^37^ for the time-stratified human landing catch (HLC) data for the field. The main reason is that the time-pattern of the mosquito HLC response in the field is driven by preferred biting times as well as environmental factors and not by mortality and successful landing (as opposed to semi-field where mosquitoes start host seeking readily because of starving and where environmental factors are well controlled). And even if the baseline time pattern of the mosquito HLC response in the field was known and the number of mosquitoes in the system could be assumed constant, no disarming or killing could be detected as the number of mosquitoes ‘in the system’ is much bigger than the number of mosquitoes affected by the intervention.

#### Analysis of field data

To estimate parameters from the field data, the Bayesian hierarchical models described in^38^ were re-parameterised coupling indoor and outdoor biting so that all parameters are estimated with one single joint posterior distribution, allowing for sample-based computations of further parameter transformations. Hence, the following statistical model is equivalent to the one presented in the supplementary methods to^38^ (see notation therein) and it was checked that inference with Stan’s^52^ NUTS algorithm^53^ yields almost identical parameter estimates:

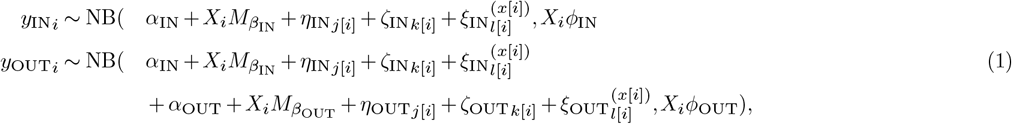

With the same second level model for both indoor and outdoor:

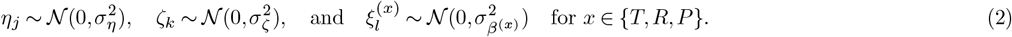

Consequently, the mean indoor and outdoor counts are given by:

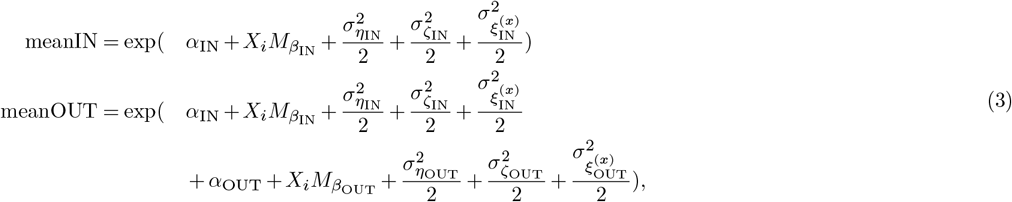

and the mean indoor and outdoor effects are given by

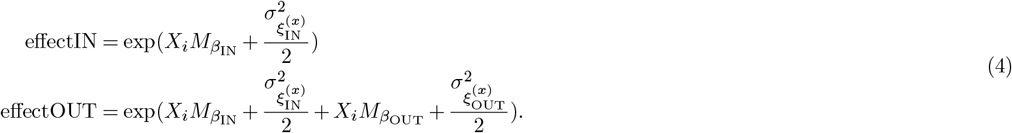

To fully account for the experimental design, a version of the model where the intervention effects were allowed to vary by house and a version where they were allowed to vary by week were fitted separately to the data and then stacked with equal weights for the final parameter estimates.

The same model was used to analyse the Suna trap catch counts in relation to outdoor biting by replacing the indoor counts with the Suna trap catch counts.

#### Repellency

The repellency effect on total mosquito host encounters for each species, comprising both indoor and outdoor landing, is given as the relative reduction of total mosquito counts (protective efficacy) being the sum of the above mean indoor and outdoor counts, yielding a formula based on the sum over indoor and outdoor effect, weighted by the mean control indoor and outdoor counts, respectively:

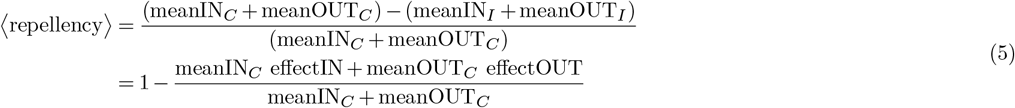

The subscript I denotes the spatial repellent or the push-pull intervention and the subscript C denotes the control. The posteriors of the repellency estimates of the spatial repellent and the push-pull system by species, including the randomly drawn samples used for the simulations, are shown in Fig. 1 of the main text.

#### Killing/disarming

The estimate of the killing/disarming effect of the spatial repellent and the push-pull intervention on *An. arabiensis* from the semi-field data (*κ*)^37^ was adjusted to the field conditions. Adjustment was made by multiplying with the ratio of the repellency (protective efficacy) estimated from the field data over the repellency estimated from the semi-field data. Note that the protective efficacy in the field trial^38^ depends on repellency only, and is actually equal to repellency, since the killing/disarming effect induced by the interventions deployed to a couple of houses is negligible given the size of the local mosquito population. Scaling by the repellency is a conservative assumption since the repellency estimated from the field data is the weighted average over indoor and outdoor repellency and thus may be low (or even non-existent) even though repellency from indoor biting may be high. However, high indoor repellency indicates that many mosquitoes may have been affected by the volatile transfluthrin while trying to enter the house and thus may have been killed or disarmed, despite a low overall repellency. The killing/disarming effect was restricted to be positive in order to be biologically meaningful, and the estimate of the disarming effect on *An. arabiensis* was therefore truncated:

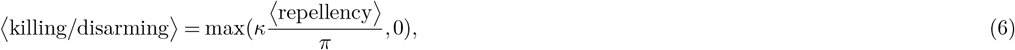

where *κ* stands for the killing/disarming parameter and *π* for the repellency parameter, as defined in^37^. However, the truncation came only into effect for *An. arabiensis* since the repellency estimated from the field data was very low for this species. The posteriors of the killing/disarming estimates of the spatial repellent and the push-pull system by species, including the randomly drawn samples used for the simulations, are shown in Fig. 1 of the main text.

#### Proportional change of indoor biting

The proportional change of the indoor biting (vs. outdoor) due to the spatial repellent, the trap or the push-pull system is given as the ratio of the indoor vs. outdoor biting proportion in the intervention arm (denoted with I) over the indoor vs. outdoor biting proportion in the control (denoted with C):

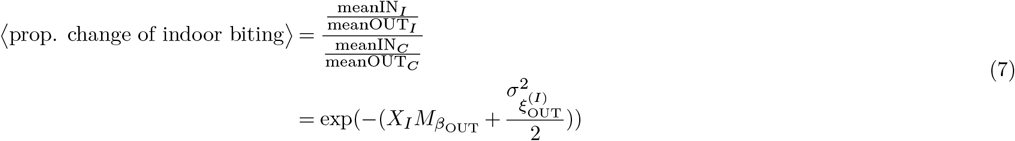

The corresponding posteriors, by species, and the randomly drawn samples used in the simulations are shown in Fig. 1 of the main text.

The change of the indoor vs. outdoor biting proportion by the interventions is incorporated into the INT parameterisation via the indoor vs. total biting proportion (*π*_*i*_ value). The indoor vs. total biting proportion under deployment of one of the interventions (*π*_*i*_^*I*^ value) is obtained by:

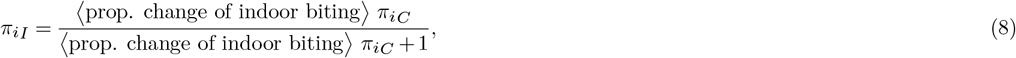

where *π*_*i*_^*C*^ denotes indoor vs. total biting proportion from the literature (see Table 3), depending on both the species and the location. The corresponding posteriors, by species and by location, and the randomly drawn samples used in the simulations are shown in Fig. S1.

#### Killing after biting

Killing after biting effect of the spatial repellent on *An. arabiensis* was estimated from the semi-field data as described in^37^. As no other data was available the same estimate was also used for the push-pull system, and for both species. The corresponding posterior and the randomly drawn samples used in the simulations are shown in Fig. 1 of the main text.

**Figure S1.**
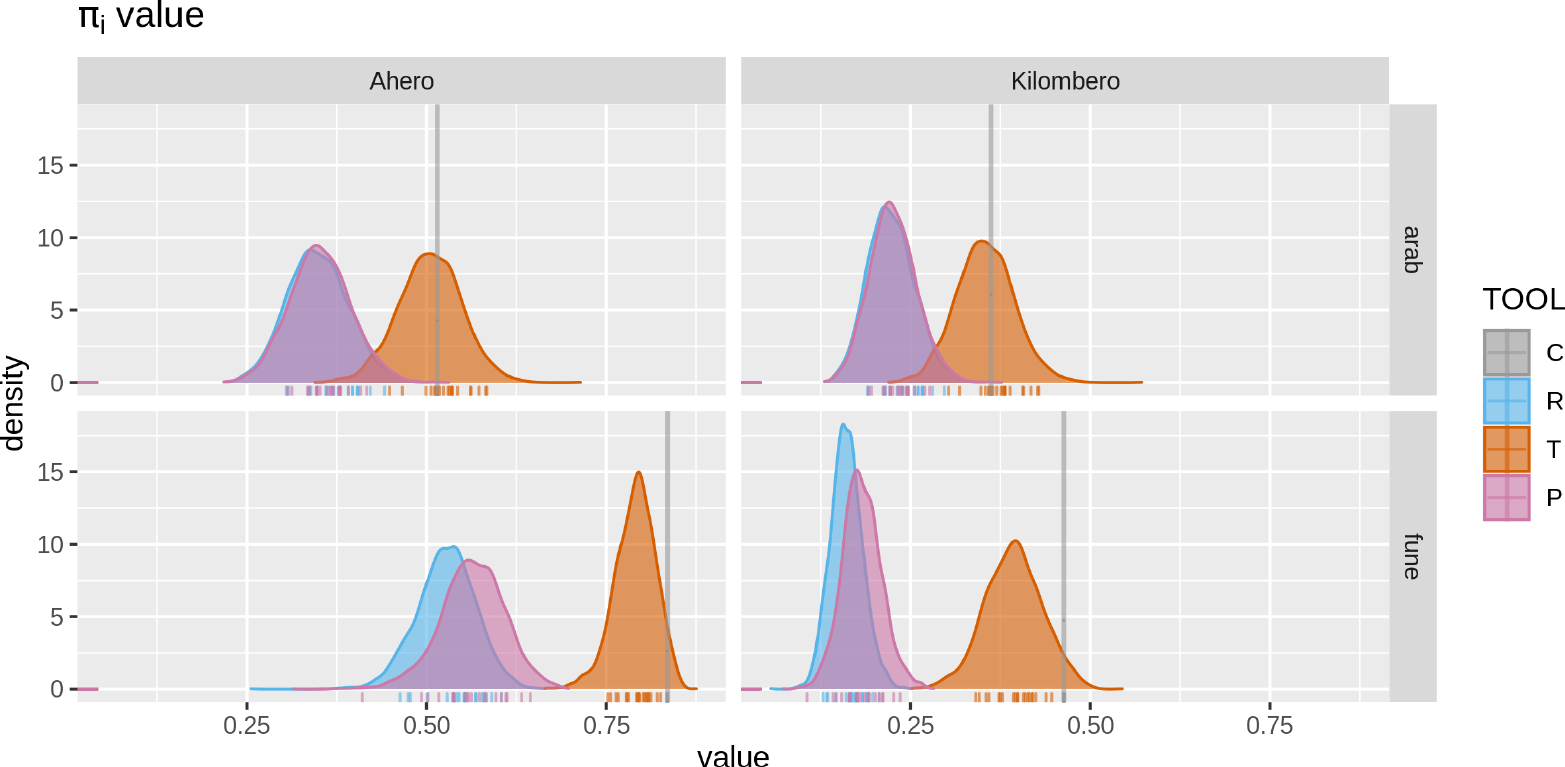
Posteriors of the indoor vs. total biting proportion (*π*_*i*_ value) under deployment of the spatial repellent (R), the trap (T) and the push-pull system (P) by species and by location, and the corresponding randomly drawn samples used in the simulations.

#### Relative trap availability

The relative availability of the Suna trap, either deployed alone or as part of the push-pull system, as compared to an unprotected human outdoors is given by:

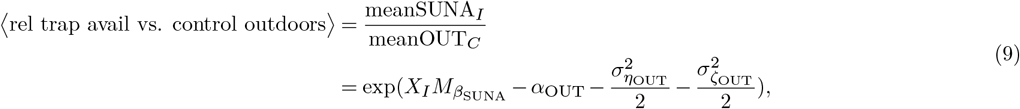

where I stand either for the trap or the push-pull intervention. The corresponding posteriors, by species, and the randomly drawn samples used in the simulations are shown in Fig. 1 of the main text.

This parameter is then multiplied with the outdoor vs total biting proportion of the given location to obtain the relative trap availability with respect to an unprotected human:

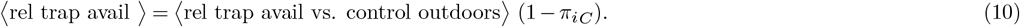

The corresponding posteriors, by species and by location, and the randomly drawn samples used in the simulations are shown in Fig. S2.

#### Implementation of intervention effects

#### Killing and disarming effect

The killing and disarming effects are implemented in OpenMalaria by adding a shadow host^37^ for each protected human host whose availability rate is given by multiplying the mean availability rate of an unprotected human hosts, determined by the adult availability rate and the age-dependent availability function applied to the given demography, with *κ*. Mosquitoes that encounter a shadow host are either killed under the killing scenario, or restart host seeking 2 days later after completing a feeding cycle under the disarming scenario.

#### Repellency and killing after biting

The repellency and the killing after biting effect of the spatial repellent and the push-pull system are implemented in OpenMalaria through the ‘general vector control’ (GVI) intervention^65^.

#### Proportional change of the indoor vs. total biting proportion and interaction with ITNs

The interaction of the spatial repellent, the trap and the push-pull system with the ITN was implemented by altering the ITN effectiveness according to the indoor vs. total biting proportion under deployment of one of the interventions (*π*_*iI*_ value), analogously to the exposition in Section for *π*_*i*_.

#### Suna trap

The Suna trap was implemented in OpenMalaria via the ‘non-human host’ intervention^65^. Since there was no functionality in OpenMalaria to set the number of non-human hosts of a given type at the time, the relative trap availability was multiplied with the given coverage level and divided by the house-hold size.

**Figure S2.**
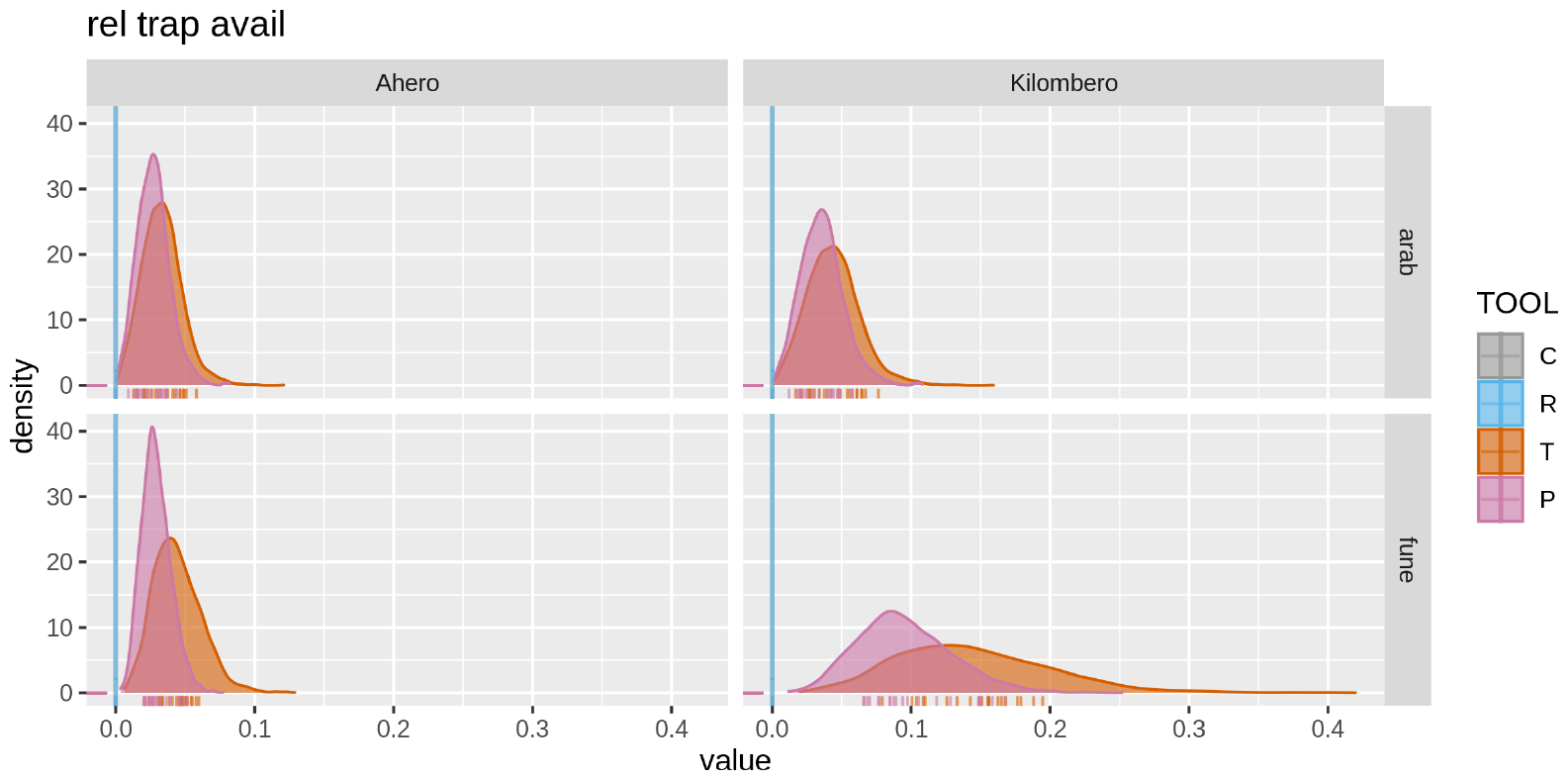
Posteriors of the relative availability of the trap as compared to an unprotected average human for the trap (T) and the push-pull system (P) by species and by location, and the corresponding randomly drawn samples used in the simulations.

#### Baseline INT paramterisation

The deployment of ITNs - including decay of insecticide concentrations, formation of holes and attrition - is implemented in OpenMalaria as explained in the Appendix to^66^, and we used the parameterisation from the corresponding electronic supplementary information, which assumes uniform effects on indoor biting of all anopheles species. The insecticidal effect decays exponentially with a half time of 1.5 year, while the net itself decays in a smooth-compact manner over about 20 years.

The parameterisation of the ITN according to^66^ is estimated from the outcomes of experimental hut studies. As suggested in^67^, we adjusted the ITN parameterisation for different indoor biting ratios (*π*_*i*_ value) by weighting the effect of the INT on the experimental hut outcomes with *π*_*i*_.

### Additional Results

#### Ahero

##### Kilombero

**Figure S3.**
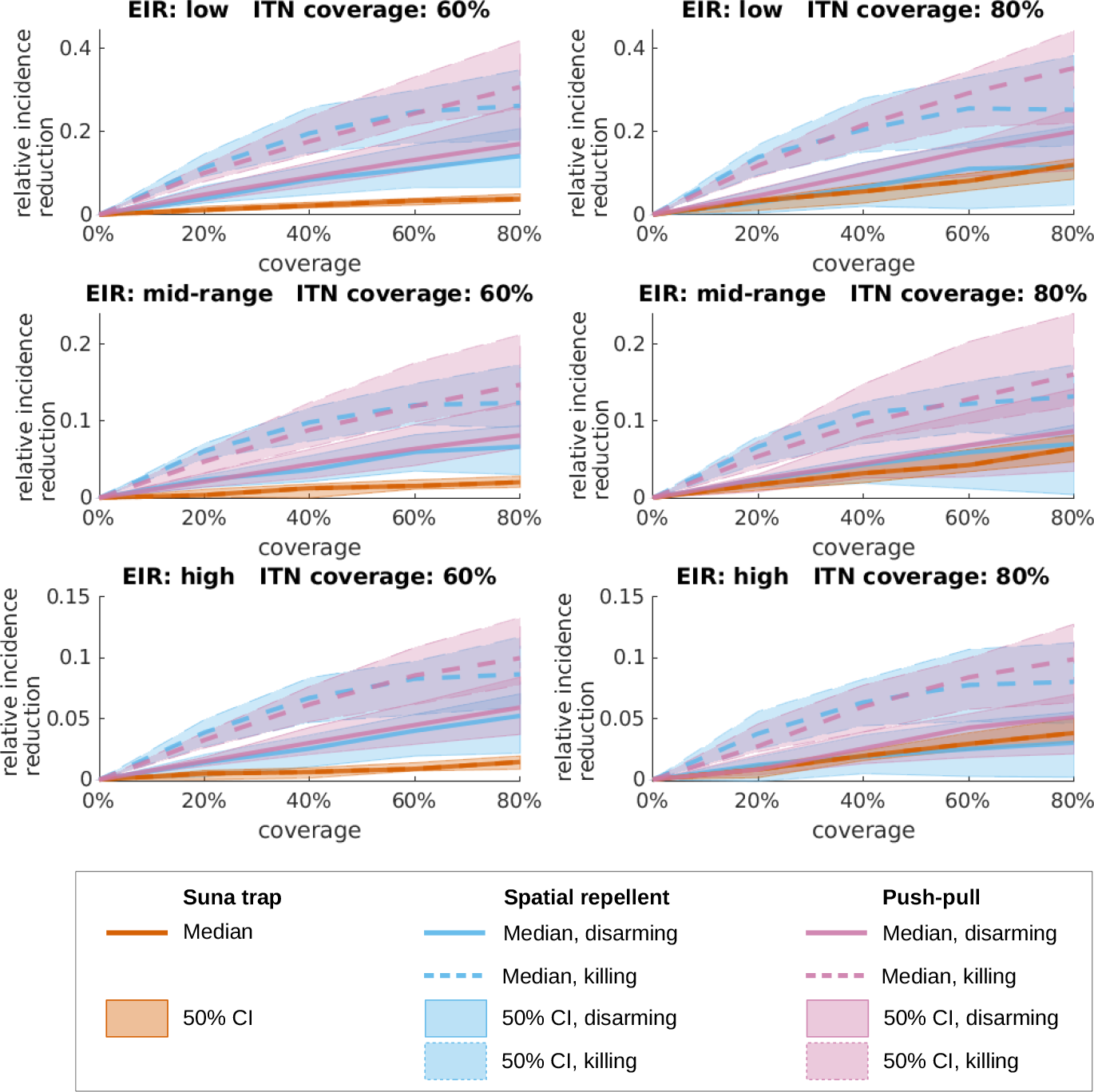
Predicted relative malaria incidence reduction when increasing use of the eave ribbon (blue), the Suna trap (red) or the combined push-pull system (purple) from 0 to 80% of the human population in Ahero. Sub-panels show different baseline assumptions with respect to transmission intensity, as measured by the entomological inoculation rate (EIR), and ITN coverage. At baseline, the low, mid-range and high transmission settings correspond to EIRs of approximately 13, 30 and 60 under the 60% ITN coverage scenario and to EIRs of approximately 8, 20 and 40 under the 80% ITN coverage scenario (Fig. S4). The interventions under consideration are allocated to people regardless of their ITN ownership (random mixing). The curves represent median predictions and the shaded areas depict 50% credible intervals (equal tailed, from 25 to 75% percentile). For the eave ribbon and the push-pull system, the mosquitoes who didn’t reattempt biting during the same night after exposure to the volatile transfluthin in the semi-field experiments, are assumed to either rest for 2 days (disarming, solid lines) or die within 24 hours (dashed lines).

**Figure S4.**
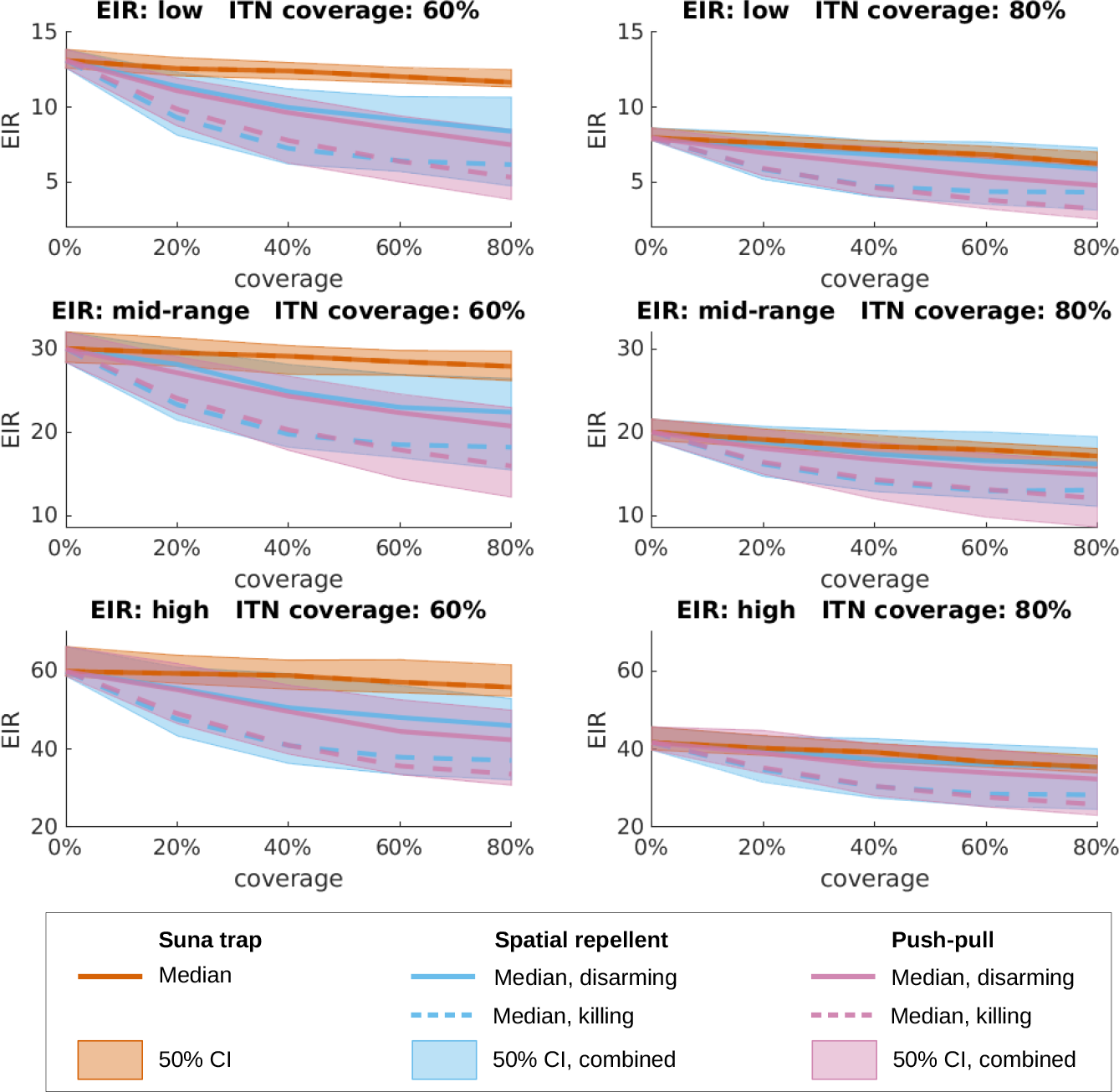
Simulated EIR. Estimates of the entomological inoculation rate (EIR) under different transmission settings in Ahero with either the spatial repellent (blue), Suna trap (red) or push-pull (purple) interventions with increasing coverage, under the assumption that the new interventions are allocated to people regardless of their ITN ownership (random mixing). Lines denote the median estimate, with solid lines standing for the disarming and dashed lines for the killing assumption for the spatial repellent and push-pull interventions. The shaded areas show 50% credible intervals (equal-tailed intervals), combining the killing and disarming scenarios for the spatial repellent and push-pull interventions (lower 25% percentile of the killing scenario to upper 75% percentile of the disarming scenario). Note that the vertical axes are aligned per row, but not across all transmission settings, and that the vertical axes do not start at 0.

**Figure S5.**
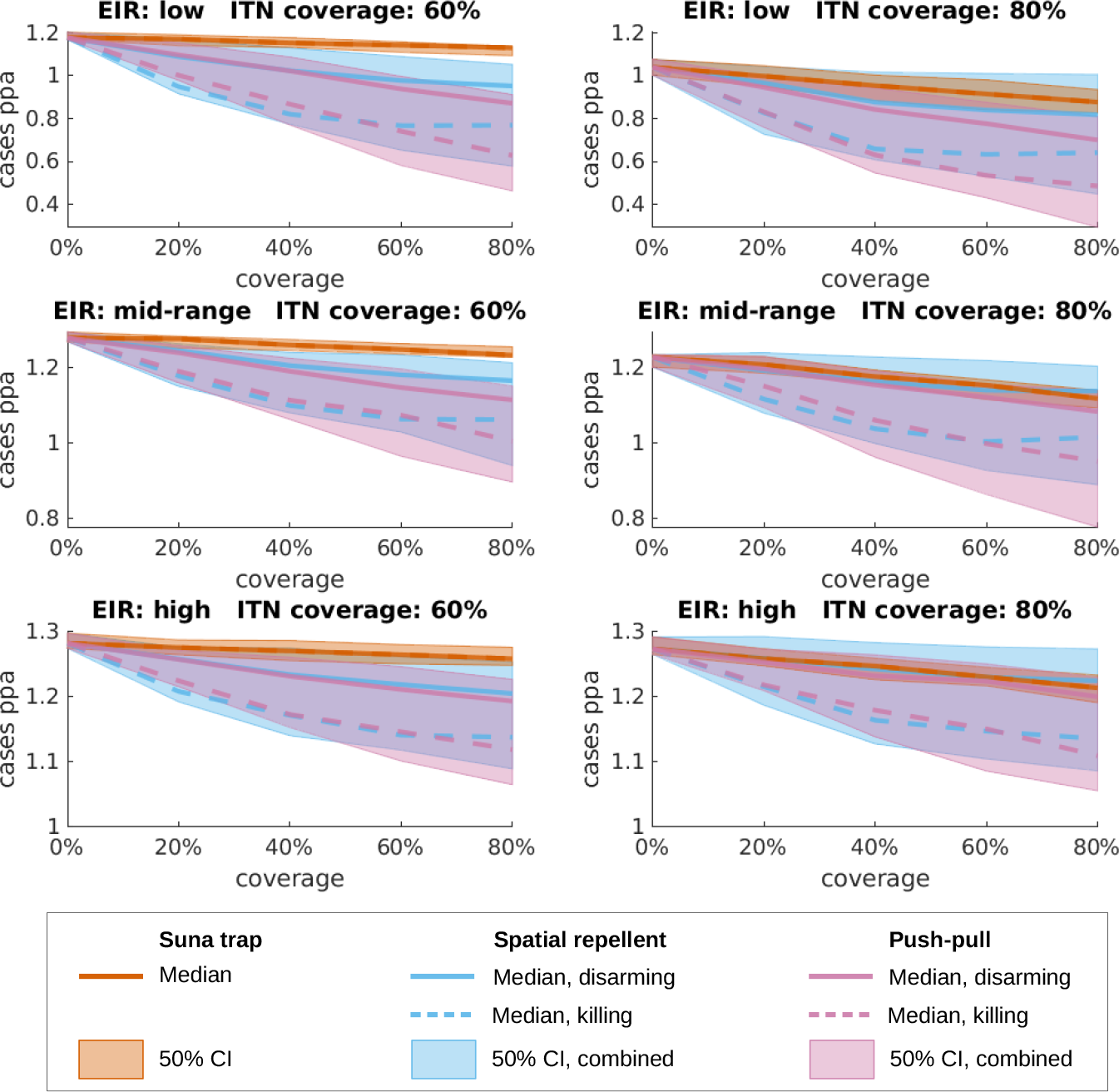
Incidence of uncomplicated malaria under assumption of increased case management coverage at baseline. Estimates of the number of uncomplicated malaria episodes per person per year (cases ppa) under different transmission settings in Ahero with increased case management coverage at baseline, with either the spatial repellent (blue), Suna trap (red) or push-pull (purple) interventions with increasing coverage, under the assumption that the new interventions are allocated to people regardless of their ITN ownership (random mixing). Lines denote the median estimate, with solid lines standing for the disarming and dashed lines for the killing assumption for the spatial repellent and push-pull interventions. The shaded areas show 50% credible intervals (equal-tailed intervals), combining the killing and disarming scenarios for the spatial repellent and push-pull interventions (lower 25% percentile of the killing scenario to upper 75% percentile of the disarming scenario). Note that the vertical axes are aligned per row, but not across all transmission settings, and that the vertical axes do not start at 0.

**Figure S6.**
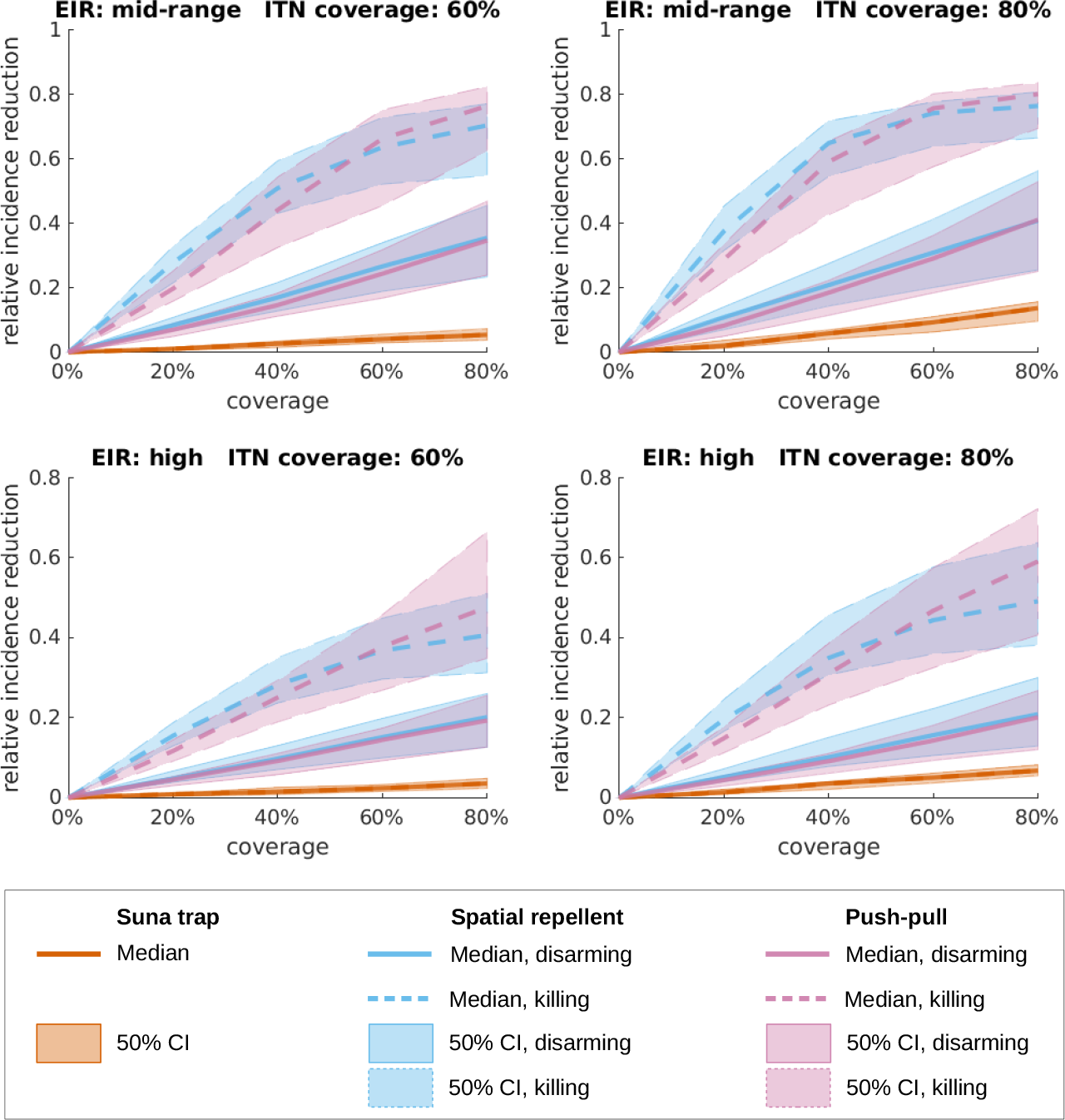
Predicted relative malaria incidence reduction when increasing use of the eave ribbon (blue), the Suna trap (red) or the combined push-pull system (purple) from 0 to 80% of the human population in Kilombero. Sub-panels show different baseline assumptions with respect to transmission intensity, as measured by the entomological inoculation rate (EIR), and ITN coverage. At baseline, the mid-range and high transmission settings corresponded to EIRs of approximately 22 and 48 under the 60% ITN coverage scenario and to EIRs of approximately 13 and 32 under the 80% ITN coverage scenario (Fig. S7). The interventions under consideration are allocated to people regardless of their ITN ownership (random mixing). The curves represent median predictions and the shaded areas depict 50% credible intervals (equal tailed, from 25 to 75% percentile). For the eave ribbon and the push-pull system, the mosquitoes who didn’t reattempt biting during the same night after exposure to the volatile transfluthin in the semi-field experiments, are assumed to either rest for 2 days (disarming, solid lines) or die within 24 hours (dashed lines).

**Figure S7.**
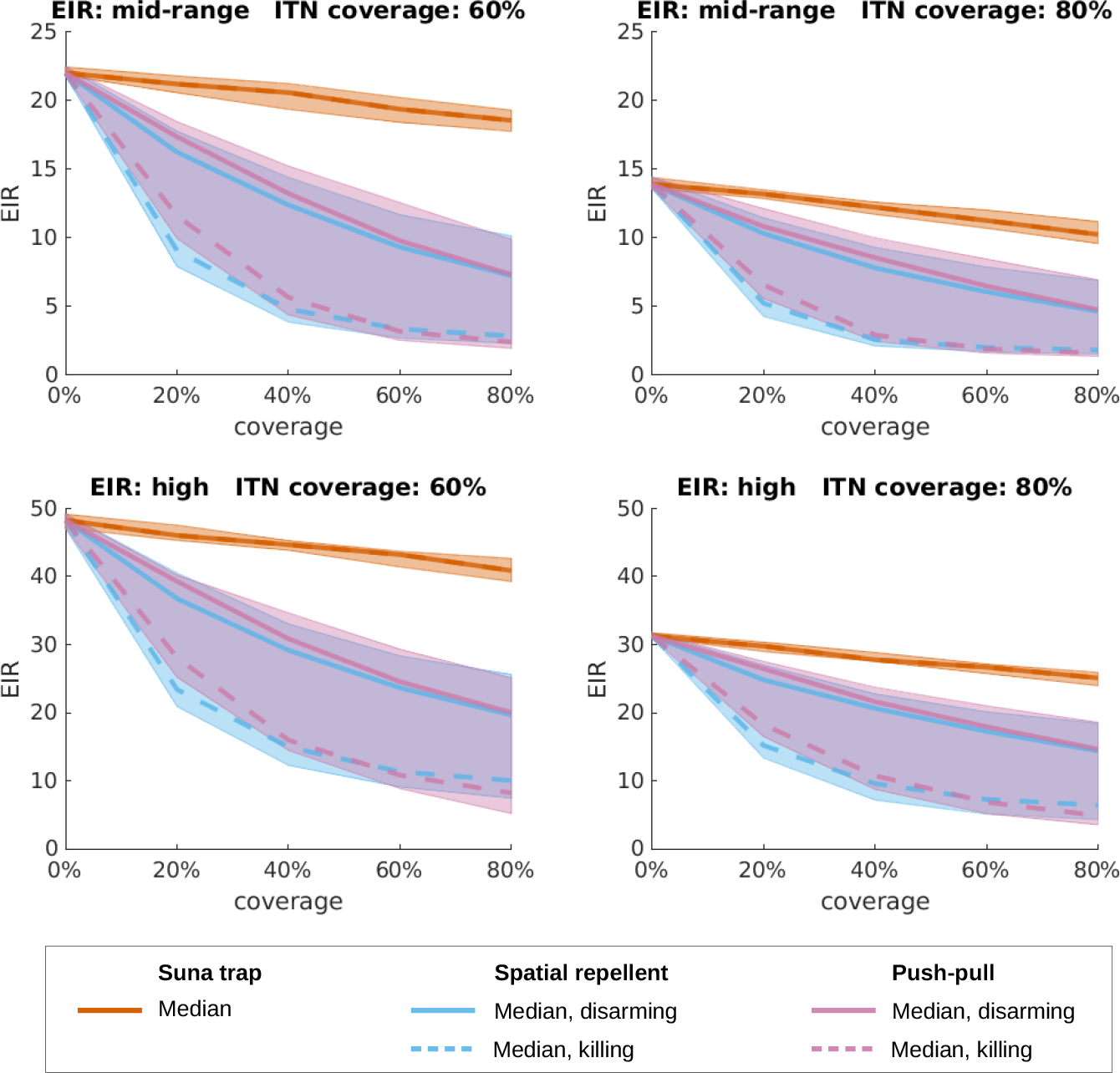
Simulated EIR. Estimates of the entomological inoculation rate (EIR) under different transmission settings in Kilombero with either the spatial repellent (blue), Suna trap (red) or push-pull (purple) interventions with increasing coverage, under the assumption that the new interventions are allocated to people regardless of their ITN ownership (random mixing). Lines denote the median estimate, with solid lines standing for the disarming and dashed lines for the killing assumption for the spatial repellent and push-pull interventions. The shaded areas show 50% credible intervals (equal-tailed intervals), combining the killing and disarming scenarios for the spatial repellent and push-pull interventions (lower 25% percentile of the killing scenario to upper 75% percentile of the disarming scenario). Note that the vertical axes are aligned per row, but not across all transmission settings, and that the vertical axes do not start at 0.

**Figure S8.**
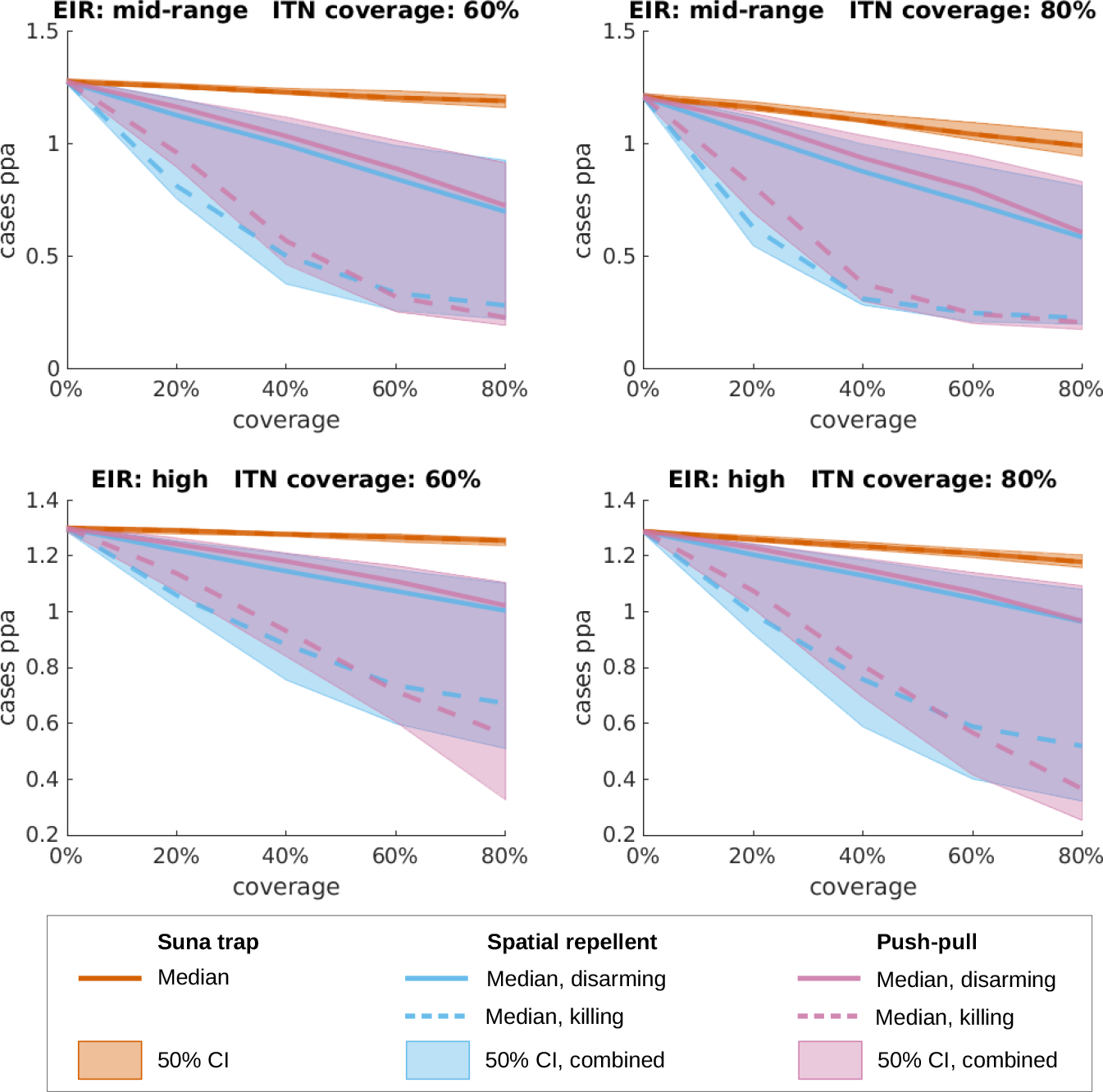
Incidence of uncomplicated malaria under assumption of increased case management coverage at baseline. Estimates of the number of uncomplicated malaria episodes per person per year (cases ppa) under different transmission settings in Kilombero with increased case management coverage at baseline, with either the spatial repellent (blue), Suna trap (red) or push-pull (purple) interventions with increasing coverage, under the assumption that the new interventions are allocated to people regardless of their ITN ownership (random mixing). Lines denote the median estimate, with solid lines standing for the disarming and dashed lines for the killing assumption for the spatial repellent and push-pull interventions. The shaded areas show 50% credible intervals (equal-tailed intervals), combining the killing and disarming scenarios for the spatial repellent and push-pull interventions (lower 25% percentile of the killing scenario to upper 75% percentile of the disarming scenario). Note that the vertical axes are aligned per row, but not across all transmission settings, and that the vertical axes do not start at 0.

